# Structured Ethical Review for Wastewater-Based Testing

**DOI:** 10.1101/2023.06.12.23291231

**Authors:** Devin A. Bowes, Amanda Darling, Erin M. Driver, Devrim Kaya, Rasha Maal-Bared, Lisa M. Lee, Kenneth Goodman, Sangeet Adhikari, Srijan Aggarwal, Aaron Bivins, Zuzana Bohrerova, Alasdair Cohen, Claire Duvallet, Rasha A. Elnimeiry, Justin M. Hutchison, Vikram Kapoor, Ishi Keenum, Fangqiong Ling, Deborah Sills, Ananda Tiwari, Peter Vikesland, Ryan Ziels, Cresten Mansfeldt

## Abstract

Wastewater-based testing (WBT) for SARS-CoV-2 has rapidly expanded over the past three years due to its ability to provide a comprehensive measurement of disease prevalence independent of clinical testing. The development and simultaneous application of the field blurred the boundary between measuring biomarkers for research activities and for pursuit of public health goals, both areas with well-established ethical frameworks. Currently, WBT practitioners do not employ a standardized ethical review process (or associated data management safeguards), introducing the potential for adverse outcomes for WBT professionals and community members. To address this deficiency, an interdisciplinary group developed a framework for a structured ethical review of WBT. The workshop employed a consensus approach to create this framework as a set of 11-questions derived from primarily public health guidance because of the common exemption of wastewater samples to human subject research considerations. This study retrospectively applied the set of questions to peer-reviewed published reports on SARS-CoV-2 monitoring campaigns covering the emergent phase of the pandemic from March 2020 to February 2022 (n=53). Overall, 43% of the responses to the questions were unable to be assessed because of lack of reported information. It is therefore hypothesized that a systematic framework would at a minimum improve the communication of key ethical considerations for the application of WBT. Consistent application of a standardized ethical review will also assist in developing an engaged practice of critically applying and updating approaches and techniques to reflect the concerns held by both those practicing and being monitored by WBT supported campaigns.

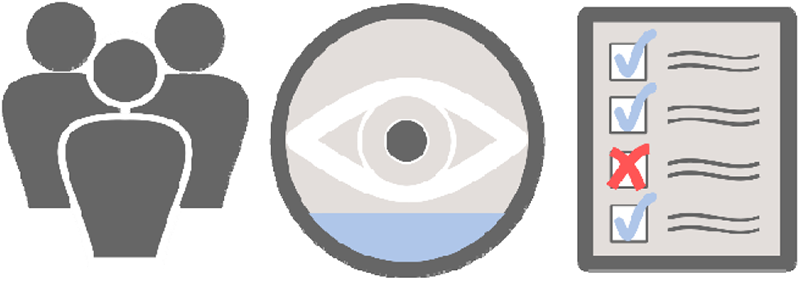

**Synopsis:** Development of a structured ethical review facilitates retrospective analysis of published studies and drafted scenarios in the context of wastewater-based testing.

## Introduction

### The Need for a Structured Ethical Review

Wastewater-based testing (WBT) describes the sampling of wastewater to support initiatives such as public health, scientific research, law enforcement, and corporate surveillance. This flexibility of WBT originates from the ability to collect wastewater samples in near real-time, at the population-level, and for a variety of analytical targets, including pathogens that cause infectious diseases. Discussing the technique of WBT often progresses rapidly to describing an application rather than exploring the concept in application-free terms^1^. For example, WBT has been used successfully to monitor enteric and respiratory pathogens, such as poliovirus and coronaviruses^2, 3^. Integrating WBT into public health surveillance is potentially less invasive, more efficient, and more inclusive than clinical testing. Inclusivity is an inherent property of the aggregated nature of wastewater, which contains target infection bioindicators (e.g., viral RNA) excreted by community members into a municipal sewer network and can capture symptomatic, asymptomatic, and pre-symptomatic carriers of infectious pathogens regardless of an individual’s access to healthcare^4^. WBT can also provide an early warning of pathogen presence within a given community, as well as detect and track circulating and novel genomic variants (e.g., for SARS-CoV-2)^5–7^. As such, the use of WBT presents as a reliable, cost-effective, objective, and rapid public health tool that complements clinical testing methods that are increasingly being adopted^8, 9^.

This application-specific consideration of WBT may too easily borrow the well-established public health ethical and legal frameworks to the analysis of a flexible tool. For example, academic researchers who used WBT to monitor pathogens were exempt from, or did not seek, standardized research ethics oversight or review because of the composite nature of the collected wastewater^10, 11^. Whereas public health departments rapidly incorporated wastewater sampling into the surveillance of SARS-CoV-2^12–14^, WBT additionally supported the research activities of academics and services offered by commercial biotechnology firms. This constellation of state, private, and academic entities is common within the operation and development of public health surveillance, with all entities performing clinical sampling. However, not all sampling translates into surveillance in support of public health, and each entity has separate legal and ethical frameworks^15^. Therefore, even when considering the ethics of a specific application of the tool, it is important to distinguish between WBT used for public health surveillance, which follows well-established professional practice and WHO guidelines^16^, and with WBT used for non-public health purposes (e.g., scientific research, law enforcement) or by non-public health entities (e.g., private entities)^17^. This distinction between testing and surveillance supports the usage of WBT throughout the text rather than the more often reported wastewater-based surveillance (WBS). “Surveillance” is a term of art, and in the context of public health, describes a method of learning about health, which is different from research or other applications of WBT. “Testing” is the technology or tool that can be used for a number of processes, including research, public health surveillance, or other types of monitoring.

When considering WBT in a more application-free context, concerns arise around data utilization and civic governance^1, 15^. Whether WBT is used for research, private monitoring, or public health surveillance, overarching ethical questions have yet to be fully explored. WBT might negatively affect privacy expectations and civil liberties at the community or even individual level^18–20^. Cases in which wastewater-derived results prompt a targeted response in a specific community could stigmatize or might, in principle, violate the privacy of the sampled community, deflating the unique feature of conducting WBT; anonymity^21, 22^. Conversely, the population at-large has a right to benefit from publicly-funded advanced surveillance technologies and the right to information about particular outbreaks so that individuals can make informed decisions about their health^15^, which ultimately requires broad and transparent communication of WBT results^23^. Generally, the usage of WBT to monitor for diseases, toxins, and terrorists threats receive broad public support across the United States ^24^. Accordingly, special attention should be paid to contextualize WBT in terms of culture and community values, the intended result of the testing efforts, and the individuals connected to the sewer conveyance network, such that results can be communicated quickly, effectively, equitably, and ethically to maintain community buy-in^25, 26^. A similar point is made by those who advocate for community-based participatory research whenever academic or government investigators seek to learn more about groups of people in a particular or specific region or place. Notably, with specific sampling place primarily occurring in fixed, publicly-owned or operated infrastructure, unique considerations arise in comparison to other public health measures surrounding the civic governance of WBT, necessitating development of a comprehensive framework that captures this multi-profession collaboration^15^.

An ethical framework applied to WBT might limit challenges that arise in contexts characterized by persistent injustice or violations of human rights. In addition, clearly defined ethical practices and processes can reduce or prevent community harm from and resistance to WBT when used for public health surveillance or scientific research purposes. The importance of clear ethical guidelines was recognized by the WHO’s general framework for ethical public health surveillance systems^16^. Thus, WBT practitioners, which include a mix of scientific experts from highly diverse disciplines and public health authorities, are increasingly called to translate this general framework into practice^27^, to both protect the public and ensure a high level of support from the community with broad social acceptance and trust^28–30^. However, as demonstrated by lack of independent review and oversight of WBT for SARS-CoV-2 monitoring, translating these guidelines into professional practice remains unstandardized and in an early phase of adoption due to those initially conducting WBT having expertise and training outside of public health practice. Additionally, as WBT continues to expand into further applications that transcend multiple disciplines^23^, such as opioid detection, monitoring campaigns should be reassessed for each new application, community, and location^31^. Therefore, interpreting the existing public health frameworks set forth by the WHO and others^15, 30, 32–34^ and translating them into a concrete, actionable, and specific framework of questions, can assist wastewater practitioners, public health officials, policy makers, utilities, and the public in interpreting the suitability of WBT.

This question-oriented framework can also encourage new entities engaging in the field to rapidly adopt best practices and can provide the tools to identify those applications that fail to align. Critically, the framework is provided as a set of questions to interrogate an application, not a set of finalized guidelines. This framework identifies concerns concretely and rapidly to enable interdisciplinary teams to engage well-established professional practices in collaboration. Additionally, this framework highlights areas that require further review rather than providing a strict protocol given that ethical issues are often easily raised, yet require contextualized analysis and continued engagement by all involved to address successfully. Finally, this standardized set of questions might also promote an ethical research culture, if adopted and upheld as an ongoing practice, and support the reputation and trust of the research field, ensuring equitable and sustainable foundations for WBT systems and community engagement^35^. For these reasons, we present a structured ethical review framework designed as a worksheet to assist in ensuring the successful, long-term, and wide-ranging implementation of WBT.

## Methods

### Design of a Structured Ethical Review

Participants contributing to the development of the structured ethical review were recruited through a public announcement at the Water Environment Federation’s Public Health and Water Conference & Wastewater Disease Surveillance Summit on March 23, 2022, as well as active social media announcements and word-of-mouth referrals. From this effort, 29 active participants were involved in the formulation of this study. Participants included representatives from a range of WBT activities, including academic researchers, public health and wastewater practitioners, and private entities working in WBT. The workshops drew upon two previously published articles describing ethical considerations of surveillance to develop the framework for a structured ethical review of the existing COVID-19 WBT literature, with the concepts of structured reviews being well-established in the creation of Institutional Review Board (IRB) processes^36^. The first selected article, written by Gary Marx (1997), emphasizes what the author calls “the new surveillance”^37^. Marx (1997) poses 29 questions in three categories (the means, the data collection context, and uses) to assess the ethics of surveillance, which the workshop then adapted to better suit WBT. The second article, by Hrudey et al. (2021), explicitly focused on ethical guidelines through 17 questions applied to SARS-CoV-2 wastewater surveillance based on a comprehensive literature review and previous WHO recommendations^32^. Hrudey et al. concluded the existing public health ethics literature fails to provide robust guidance for WBT practitioners.

To apply and add to these previous works, the participants developed the structured ethical review framework using a workshop approach. Prior to the workshop, each participant reviewed these two articles on ethics of public health surveillance^32, 37^ and drafted concise descriptions for three levels of ethical sufficiency for each of the categories posed by Marx (1997) and Hrudey et al. (2021) applied to WBT activities. The three levels of ethical sufficiency were as follows: 0 - minimal review required (no ethical concerns); 1 - review suggested (limited ethical concerns); and 2 - review strongly suggested (broad ethical concerns). These levels were selected to prioritize ethical reviews and communication efforts of those designing and operating surveillance programs. Thereafter, each participant independently filled their brief written descriptions within a shared document for each previously identified category of ethical consideration identified in the two articles. During the virtual workshop, participants reviewed the responses and identified the guidelines and questions that warranted further discussion; the output thus identified consensus descriptions of ethical sufficiency rankings. After the workshop, each participant adopted a category and prepared a final draft description for the three levels of ethical sufficiency into a table (Supplemental Table 1). All co-authors then reviewed the table, and revisions were made until a consensus final draft was reached, with duplicate categories merged, but all others preserved from Marx (1997) and Hrudey et al. (2021). The final, fully consolidated framework comprises 37 categories of ethical consideration for WBT, with three ordinal ranking descriptions within each category (Supplemental Table 1). These categories broadly represent key considerations in community engagement, equality, establishment of a new precedent, and data integrity. With the large size of the framework, the set was further refined, with the participants being asked to list ten essential questions to include. In total, 16 participants provided a score, and those categories receiving over 9 votes or higher were included in the final framework (Table 1). In essence, the developed structured ethical review provides a set of questions that enables users to provide a score (higher the score the greater the ethical concern) when considering a WBT application.

**Table 1.** Consolidated framework for a structured ethical review to assess potential adverse outcomes of WBT efforts. Specific categories originated from either Marx 1997 [*] or Hrudey et al., 2021 [†]. The top 11 categories based on the internal voting are presented here, the full framework is presented as Supplemental Table 1, available at DOI:10.17632/2xkfkcsxx8.1.

| Category | 0 - Minimal Review Required | 1 - Review Suggested | 2 - Review Strongly Suggested |
| --- | --- | --- | --- |
| **Legitimacy:** Are surveillance data collected only for a legitimate public health purpose?† | Data support public health agents for public health measures | Data support non-public health agents for public health measures | Data support non-public health agents (or public health agents acting outside of public health) for non-public health purposes |
| **Unfair Disadvantage:** Is the information used in such a way as to cause unwarranted harm or disadvantage to its subject?* | WBT is implemented with clear scope, oversight, and decision-making procedures including procedures for response to wastewater data with policies for follow-up clinical testing applicable to full communities | Unintentionally subjecting specific areas to the possibility of a disruptive intervention (lockdowns, quarantines, isolations of entire areas without any process for identifying and isolating relevant individuals; mandatory testing of individuals in response to wastewater data) whereas excluding others from the same level of surveillance scrutiny and response | Intentionally subjecting specific areas to the possibility of a disruptive intervention (lockdowns, quarantines, isolations of entire areas without any process for identifying and isolating relevant individuals; mandatory testing of individuals in response to wastewater data) whereas excluding others from the same level of surveillance scrutiny and response |
| **Data Stewardship and Protection:** Is the data properly maintained to protect those monitored?* | Data managed per requirements of community monitored | Data managed per requirements of funding agency | Data management plan absent |
| **Creation of Unwanted Precedents:** Is it likely to create precedents that will lead to its application in undesirable ways?* | Analysis for individual identification prohibited; explorations outside of agreed-upon community scope is explicitly prohibited | No positional statement regarding individual identification; No discussion of future research is discussed | Explicitly for identification of individuals or otherwise unethical applications |
| **Awareness:** Are individuals informed they are being monitored and why?* | Representative(s) of the monitoring campaign are in a cycle of continued community outreach and engagement during WBT and over the clearly defined reporting period providing contextualization of the scope and intent to minimize misrepresentation or misuse; those monitoring capture the questions from the community rather than the wastewater utility operators | Duration, scope, and intent is communicated and disseminated in a passive manner without contextualization or engagement OR communicated to a single representative of the community; the wastewater utility operators respond to increased inquiries but have access to those collecting the data to direct inquiries | No direct communication of the duration, scope, and intent to the monitored community members; the burden of communication falls solely on the third-party wastewater utility operators rather than the data collectors |
| **External Data Sharing:** Is the public health surveillance data shared with other public health agencies when addressing a public health need?† | Collected WBT-supported public health data is shared freely with/between public health agencies when a public health need presents or persists | Collected WBT-supported public health data only partially shared with/between public health agencies when a public health need presents or persists | No data is shared when a public health need presents or persists |
| **Public Decision-Making:** Was the decision to use WBT in surveillance arrived at through some public discussion and decision-making process?* | Surveillance program was designed in a public manner (e.g., review by elected officials and public through town halls for initial implementation and continued operation) with a good-faith effort to reach both those receptive or resistant to the objectives of public health | Program does not receive formal public authorization but is broadly supported by the public (as informed by representative public surveys) | Program does not receive formal public authorization and is not supported by the public (as informed by representative public surveys) |
| **Right of Inspection:** Are people aware of the findings of WBT supported surveillance and how they were created?* | Representative(s) of the monitoring campaign are in a cycle of continued community outreach and engagement during the sample collection and reporting period providing contextualization of the collected data to minimize misrepresentation or misuse (e.g., updating an annotated and agreed-upon internet-accessible dashboard; timely and routine public town-halls or open seminars; direct mailing to surveyed individuals) | The collected data is communicated and disseminated in a passive manner without contextualization or engagement OR communicated to a single representative of the community | No direct communication of the collected data to the monitored community members |
| **Equality-inequality:** Is WBT broadly applied to all or only those able to resist?* | Entire community is monitored (e.g., treatment plant; jail sampling that monitors the effluent of the whole jail including staff and inmates) | Representative coverage is achieved (e.g., manhole sampling, but ensuring that demographics of surveilled communities are representative of the entire city; jail sampling that has sites for staff and inmates separately) | Only protected-class communities are monitored (e.g., manhole sampling that surveils only low-GDP per capita areas; jail sampling that only surveils inmates) |
| **Community Values:** Are the values and concerns of the communities taken into account in planning, implementing, and using data from surveillance?† | Representative of the monitoring campaign are in a cycle of continued community outreach and engagement during the planning and implementing period to address the concerns and support the values of the community | Representative of the monitoring campaign are engaged during the planning period only to address the concerns and support the values of the community | No direct involvement of the monitored community members |
| **Consequence of Inaction:** What are the consequences of taking no surveillance action?* | Mortality, morbidity, or other adverse effects is imposed on community by lack of surveillance | The surveillance data does not minimize adverse effects to the community | The community benefits by not being surveyed |

### Inclusion Criteria for Published Studies Considered within the Application of the Structured Ethical Review

The initial collection of studies was obtained by searching for articles containing the keyword phrases “SARS-CoV-2” and “wastewater,” yielding a total of 5,632 articles as of February 2022. To focus on sampling strategies that potentially challenge individuals’ assumptions of privacy, further filtering was performed to include studies reporting on near-building and/or within-sewer monitoring. This involved incorporating additional modifiers such as “campus,” “nursing home not campus,” “prison not campus,” “hospital and wastewater-based and building not campus,” and “neighborhood not campus.” Consequently, the modified search categories returned 1204, 240, 119, 201, and 313 articles, respectively. Studies were excluded if they did not mention monitoring within a sewer collection system at the neighborhood or building-level scale or if they were not yet published as peer-reviewed articles. This strategy was used to narrow the database into a representative and manageable sub-selection of WBT related literature covering the early phase of the pandemic as a case study. The goal here was to emphasize the utility of the ethical framework applied to a coherent set of WBT applications rather than defining the best practices for a given application. Therefore, this structured review approach could be applied in the future to private residential settings^38^ or at wastewater treatment plant scales^39^, but both areas were excluded from current consideration. A note on private settings: If one accepts the utility of a robust ethics analysis to improve practice and foster trustworthiness, it should not matter if an educational institution, for instance, is public or private. The analysis of human biological material and data will always raise ethical issues, and it is unacceptable to suggest that private institutions ought adhere to different scientific or ethical standards.

After applying these filters, 25 near-building and 21 within-sewer SARS-CoV-2 WBT studies were identified for review. The inclusion of 7 additional articles published after the initial search that were previously in preprint status increased the number of studies included in this structured ethical review to 53 (56 publications were represented by these studies, with 3 having two publications describing the same monitoring campaign and were considered in concert). While published research articles inherently connote research use of WBT, the selected articles had potential direct applications to public health, facilitating their use in the evaluation of the structured ethical review framework.

Papers were divided across participants for review, with overlapping assignments provided. Overall, 79 reviews were completed by answering all 37 questions based on the reviewers’ analysis of the presented text alone. Conflicting assignments were evaluated, but were left unchanged, resulting in an average score for those papers. The reviews were then consolidated into a summary data frame for broad comparisons between the responses to individual questions

## Results and Discussion

### Reconsidering SARS-CoV-2: Application of the structured ethical review

The main goal of this study was to develop a framework to assist in identifying gaps in ethical considerations of WBT as a tool. This was accomplished by first developing and then demonstrating the framework by applying the structured ethical review framework to previously reported SARS-CoV-2 WBT studies (Figure 1). Application of the structured ethical review framework highlighted that throughout the published articles reviewed in this work, gaps in information were observed in both the manuscript and supplemental information (1247 of 2923 answers provided (43%) indicated “Not Recorded in the Main Text”). This absence may be the result of the inclusion of studies predominantly reported by non-public health agents (e.g., academics, researchers). For example, when compared to clinical research in which detailed informed consent (i.e., voluntariness, information disclosure, decision-making capacity, and communication of results) is required^40^, few WBT studies articulated whether consent was obtained from the studied community, whether those results were communicated back to the community, and/or whether those results were used with clear public health objectives and outcomes. It is important to note that WBT can be applied as a public health surveillance tool that would then operate within existing legal and regulatory frameworks. Within these specific applications, consent for the collection and testing of wastewater samples may not be required from individuals when a pooled sample is being analyzed, as this is considered to be part of routine public health surveillance^41^. However, there may be legal and ethical considerations around the use of the WBT data collected, for example, in research applications, and these should be addressed by relevant authorities and other frameworks. Broadly, the absence of reported information for evaluating community engagement and data safeguards reflects the current lack of a standardized ethical framework for WBT campaigns.

**Figure 1.**
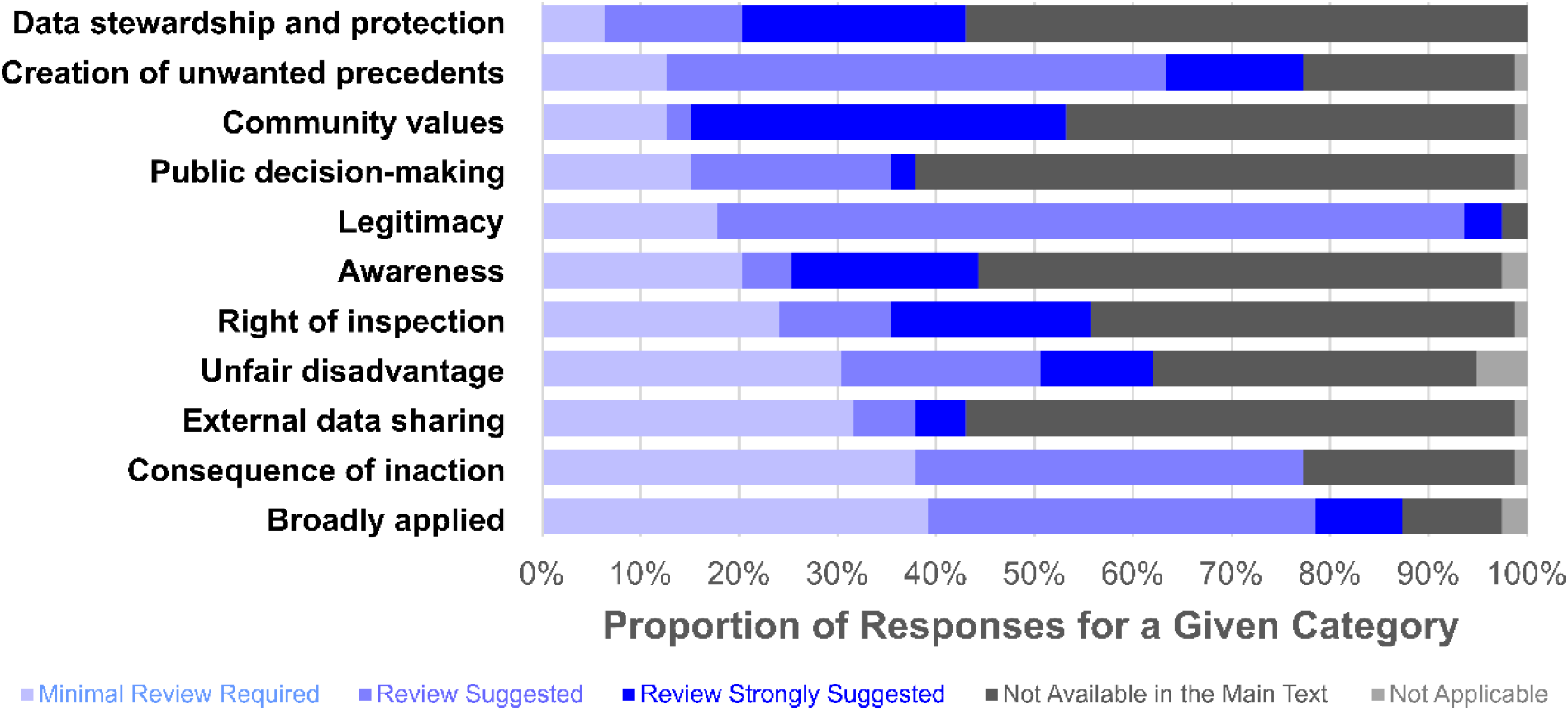
The distribution of assigned flags (“minimal review required”, “review suggested”, “review strongly suggested”, “not available in the main text”, “not applicable”) for the top 11 categories in the structured ethical review, represented as a fraction percent of all publications analyzed (n=53) with multiple reviewers providing reports for select individual studies, resulting in more reviews than studies (n=79) ^12, 42–46, 50–99^. In total, 56 publications were represented by these studies, with 3 having two publications describing the same monitoring campaign and were considered in concert. The categories are sorted by ascending proportion of “minimal review required”.

This exemption from reporting and lack of oversight may also be due to a lack of clarity for the motivation of the WBT study. For example, these articles could have been originally motivated by a research interest with results that prompted a public health action. Conversely, authors may have retrospectively published results from a public health surveillance system. In the former case, authors may have assumed the research study was exempted from IRB approval given the composite nature of the sample, which is believed to prevent the ability to identify specific individuals in a given sewer catchment^42^. In certain cases, authors referenced IRB approvals for utilizing individual case data^43^, but in most cases, WBT sample data itself was determined to be exempt from IRB oversight. In select cases, ambiguity surrounded whether the IRBs themselves arrived at these determinations or the researchers, a key consideration given that these oversight bodies are responsible for conforming to federal regulations.

However, the data collected from WBT were sometimes used for direct public health interventions, a usage of WBT that aligns with public health surveillance and that is distinct from research activities governed by IRB protocols. For example, other studies (mainly dormitory and hospital surveys) reported on positive SARS-CoV-2 detection in wastewater, triggering mandatory clinical or individualized testing from which infected individuals were identified ^43–46^ . In contrast, WBT of a cargo ship specifically explored border protection against infected seafarers as a potential public health intervention^47^, a more intervention-driven approach than that described as a complementary monitoring tool for airplanes^48^. Ultimately, the study did not support WBT in the cargo ship application, notably because individualized testing was already deployed. This highlights a key finding: the motivation for the use of WBT during the early phase of the pandemic was blurred between research and public health surveillance.

This dual nature of early monitoring campaigns complicates their interpretation. For example, if WBT was conducted as part of a public health surveillance effort, informing subsequent action where individuals were identified through individualized testing regimes (e.g., isolation), then IRB oversight would be unnecessary as this use of WBT would fall under ethical guidelines for public health surveillance. This determination that IRB is not required rests on WBT being used as a tool for public health surveillance conducted by a public health authority, which is explicitly excluded from the US IRB regulations (Subpart A of 45 CFR Part 46)^49^. However, if the wastewater data itself triggered direct public health outcomes without subsequent independent surveillance, such as clinical testing confirmation during the early phase of the pandemic, or if the subsequent clinical testing had the additional goal to produce generalizable knowledge, then IRB oversight is likely needed because both of these areas include research components during the development phase. This requirement results from either using non-clinical data, that at the time was relatively new to the public health field, for a direct health intervention; or using clinical data for a research purpose rather than public health surveillance (see Categories 4, 10, & 28 in Supplemental Table 1). Notably, none of the 53 studies reviewed with the proposed structured ethical review were flagged by their institutions for IRB approval for the WBT portion alone, suggesting that IRB approval may have been granted to other types of ongoing research such as individualized saliva testing when needed and used in combination with these programs. In compliance with the IRB regulations, studies that pair WBT with public health surveillance should seek IRB review to address the research aspects of the work.

In contrast to the ambiguity surrounding IRB review reporting, the WBT community has already taken to reporting on other ethical areas including the scale of testing (e.g., wastewater treatment plants [WWTPs], building-level), the identifiability of populations represented, the presence/absence of validated QA/QC workflows, and the need for clear statements of goals supporting public health. However, gaps in presenting ethical considerations were found with respect to stakeholder participation in the development and deployment of WBT efforts as well as data and sample management. Only 5 of 53 studies clearly identified a data management plan, with no study combining an additional communication or engagement plan. Elements that were considered when screening for a communication or engagement plan included statements surrounding how or whether the public or public representatives were engaged in the development, deployment, or future applications of WBT; the surveilled parties knew their rights or were given the right to challenge, express grievances, or seek redress; and if the potential risks and/or benefits were outlined in detail to these populations or third parties. It is possible that some studies developed communication and engagement plans that were not explicitly reported in the published research, and we acknowledge this limitation in our review. Further, wastewater data security, handling, and subsequent use in and outside the scope of the project (including the fate of remaining samples) were largely neither acknowledged nor discussed. Details on data ownership, security, management plans, and dissemination were generally absent, but key elements were provided in select studies^44, 51, 82, 96, 98^. Several elements that require more explicit elaboration include how data were reported during the operation of the campaign, by what mechanism, at what frequency (weekly, bi-weekly, etc.), and how thresholds of concern were established in terms of viral titers.

In the case of campus- or dormitory-wide testing at colleges and universities, authors provided more information for these programs when compared to studies for large sewersheds, for instance at the level of a WWTP. Specifically, these studies detailed follow-up procedures for wastewater samples that resulted in positive detection of the virus (e.g., lockdowns, contact tracing), stakeholder engagement, and data dissemination plans. Likely, this higher level of detail resulted from building-level monitoring programs intentionally designed to use WBT to assist clinical testing and quarantine procedures. However, of all papers reviewed, only a few noted a process for obtaining consent from the studied populations^45, 50, 63, 78, 80, 84, 90, 99^.

### Learning from SARS-CoV-2: Sustaining Future Applications of WBS

The structured ethical review aims to provide a framework to assess new areas or targets of monitoring and to document evolving ethical applications of WBT when uniting technical innovation, community engagement, and professional collaboration^100^. For instance, the unique characteristics of an orthopoxvirus outbreak, which differs in transmission classification and carries higher pre-existing stigma, present additional ethical considerations within WBT^101^. Future applications will present new and unique challenges to the ethical application of WBT as a tool, promoting a continuous evaluation of the structured ethical review adopted here. Therefore, establishing, applying, and updating this structured ethical review over time will result in a transparent record of our understanding of the best ethical practices when applying WBT within and beyond public health surveillance.^99^

Future WBT structured ethical reviews may need to consider human-specific, rather than just pathogen-specific, target biomolecules. Early within the development of this structured ethical review, it was established that the purview of recovering human genetic material was not the main focus of this tool given that considerations for targeted human monitoring are already developed in the biomedical research community and remain a bioethics concern beyond that of WBT^102^. However, with the technical capabilities of WBT advancing, the application of next-generation sequencing tools to collect personally identifiable health information opens unique and potentially community-desired possibilities^103^. Although relatively few papers used sequencing techniques and, when applied, were exercised only for detecting variants of SARS-CoV-2 in which human-specific DNA is masked from publicly-posted samples, there is an evident lack of guidelines when analyzing complete genetic data recovered from wastewater^18^. Notably, previous applications successfully applied more targeted approaches to screen for human mitochondrial sequences within wastewater as a population biomarker highlighting outside of sequencing technologies^104^. This necessitates a better understanding of the views and tolerances of those conducting targeted human-DNA testing, those using the data, and persons whose samples are being tested^105^, potentially informing the evolution and revision of the categories within the structured ethical review.

Importantly, this framework does not define which applications of WBT are ethically appropriate and which are not; it is simply a tool to guide the development and evolution of WBT campaigns by highlighting aspects which may require additional ethical review. In an ideal world, all WBT campaigns would receive ratings of “minimal review required” for all categories, but in practice this will almost certainly never be the case. Applying WBT requires trade-offs to maximize benefit and minimize harms. For example, waiting to implement WBT for a novel pathogen until all ethical concerns are fully resolved may hinder progress in public health surveillance to identify new outbreaks and intervene in their early stages. Different categories within the framework can also be in tension with one another, which is common for large frameworks and, thus, is expected. Additionally, the interpretation of each category will change accordingly as sampling campaigns are run by or analyzed through the viewpoint of researchers, governmental public health agencies, or private entities.

The role and utility of ethics analysis in public health, research, and clinical practice is rarely to give approvals or disapprovals of inherently challenging issues and conflicts. It is, rather, to inform an already complex environment or problem by making clear salient values – some of which might be in conflict – and help practitioners weigh and apply these values. How, for instance, ought scientists balance a duty to protect privacy with an equally compelling right to benefit from science? The challenge is lensed when reasonable people disagree about an appropriate course of action. In such cases, the existence of an objective and transparent ethics process can guide both investigators and communities such that whatever action is decided, there will be a mutual understanding of the cause. It is our view and recommendation that WBT and the communities it serves will improve with the adoption of structured ethics reviews.

As WBT expands and new practitioners and researchers enter the field, this structured ethical review framework will provide education and guidance to promote best practices ^106^. Given the length of the full, structured ethical review in terms of the number of categories, further development surrounding the ease-of-use is required to ensure wide-scale adoption of the structured ethical review framework by the WBT community. This adaptive, iterative approach to improving the framework is critical for managing this technology in the future, to safeguard the well-being of those under surveillance, and communicate robust ethical guidelines to protect the intended applications. Therefore, we strongly recommend the implementation of the structured ethical review by all those involved in both ongoing and future WBT campaigns to promote activities supporting ethical best practices.

## Data Availability

All data produced in the present study are available upon reasonable request to the authors.

## Acknowledgements

We thank Dr. Kyle Bibby (University of Notre Dame), Dr. Steve Hrudey (University of Alberta), William Johnson (University of Colorado Boulder), Dr. Todd Schenk (Virginia Tech), Dr. Scott Meschke (University of Washington), Dr. Scott Olesen (Biobot Analytics), and Dr. Rachel Poretsky (University of Illinois Chicago) for their participation in the workshops. We also thank Dr. Stefan Wuertz (University of California Davis) for feedback on an initial outline of this work. This work was supported in part by the National Institute of General Medical Sciences (NIGMS) under award numbers P20GM113117 for the participation of Justin Hutchison and by the National Science Foundation (NSF) under award number NSF 2047470 for the participation of Fangqiong Ling.

## REFERENCES

1. Doorn, N. Wastewater Research and Surveillance: An Ethical Exploration. Environ. Sci. Water Res. Technol. 2022, 8 (11), 2431–2438. https://doi.org/10.1039/D2EW00127F.

2. Ivanova, O. E.; Yarmolskaya, M. S.; Eremeeva, T. P.; Babkina, G. M.; Baykova, O. Y.; Akhmadishina, L. V.; Krasota, A. Y.; Kozlovskaya, L. I.; Lukashev, A. N. Environmental Surveillance for Poliovirus and Other Enteroviruses: Long-Term Experience in Moscow, Russian Federation, 2004–2017. Viruses 2019, 11 (5), 424. https://doi.org/10.3390/v11050424.

3. Más Lago, P.; Gary, H. E.; Pérez, L. S.; Cáceres, V.; Olivera, J. B.; Puentes, R. P.; Corredor, M. B.; Jímenez, P.; Pallansch, M. A.; Cruz, R. G. Poliovirus Detection in Wastewater and Stools Following an Immunization Campaign in Havana, Cuba. Int. J. Epidemiol. 2003, 32 (5), 772–777. https://doi.org/10.1093/ije/dyg185.

4. Chavarria-Miró, G.; Anfruns-Estrada, E.; Martínez-Velázquez, A.; Vázquez-Portero, M.; Guix, S.; Paraira, M.; Galofré, B.; Sánchez, G.; Pintó, R. M.; Bosch, A. Time Evolution of Severe Acute Respiratory Syndrome Coronavirus 2 (SARS-CoV-2) in Wastewater during the First Pandemic Wave of COVID-19 in the Metropolitan Area of Barcelona, Spain. Appl. Environ. Microbiol. 2021, 87 (7), e02750–20. https://doi.org/10.1128/AEM.02750-20.

5. Crits-Christoph, A.; Kantor, R. S.; Olm, M. R.; Whitney, O. N.; Al-Shayeb, B.; Lou, Y. C.; Flamholz, A.; Kennedy, L. C.; Greenwald, H.; Hinkle, A.; Hetzel, J.; Spitzer, S.; Koble, J.; Tan, A.; Hyde, F.; Schroth, G.; Kuersten, S.; Banfield, J. F.; Nelson, K. L. Genome Sequencing of Sewage Detects Regionally Prevalent SARS-CoV-2 Variants. mBio 2021, 12 (1), e02703–20. https://doi.org/10.1128/mBio.02703-20.

6. Lin, X.; Glier, M.; Kuchinski, K.; Ross-Van Mierlo, T.; McVea, D.; Tyson, J. R.; Prystajecky, N.; Ziels, R. M. Assessing Multiplex Tiling PCR Sequencing Approaches for Detecting Genomic Variants of SARS-CoV-2 in Municipal Wastewater. mSystems 2021, 6 (5), e01068–21. https://doi.org/10.1128/mSystems.01068-21.

7. Sutton, M.; Radniecki, T. S.; Kaya, D.; Alegre, D.; Geniza, M.; Girard, A.-M.; Carter, K.; Dasenko, M.; Sanders, J. L.; Cieslak, P. R.; Kelly, C.; Tyler, B. M. Detection of SARS-CoV-2 B.1.351 (Beta) Variant through Wastewater Surveillance before Case Detection in a Community, Oregon, USA. Emerg. Infect. Dis. 2022, 28 (6), 1101–1109. https://doi.org/10.3201/eid2806.211821.

8. Karthikeyan, S.; Ronquillo, N.; Belda-Ferre, P.; Alvarado, D.; Javidi, T.; Longhurst, C. A.; Knight, R. High-Throughput Wastewater SARS-CoV-2 Detection Enables Forecasting of Community Infection Dynamics in San Diego County. mSystems 2021, 6 (2), e00045–21. https://doi.org/10.1128/mSystems.00045-21.

9. Hart, O. E.; Halden, R. U. Computational Analysis of SARS-CoV-2/COVID-19 Surveillance by Wastewater-Based Epidemiology Locally and Globally: Feasibility, Economy, Opportunities and Challenges. Sci. Total Environ. 2020, 730, 138875. https://doi.org/10.1016/j.scitotenv.2020.138875.

10. Wolfe, M. K. Invited Perspective: The Promise of Wastewater Monitoring for Infectious Disease Surveillance. Environ. Health Perspect. 2022, 130 (5), 051302. https://doi.org/10.1289/EHP11151.

11. Verovšek, T.; Krizman-Matasic, I.; Heath, D.; Heath, E. Site- and Event-Specific Wastewater-Based Epidemiology: Current Status and Future Perspectives. *Trends Environ*. Anal. Chem. 2020, 28, e00105. https://doi.org/10.1016/j.teac.2020.e00105.

12. Layton, B.; Kaya, D.; Kelly, C.; Williamson, K.; Bachhuber, S.; Banwarth, P.; Bethel, J.; Carter, K.; Dalziel, B.; Dasenko, M.; Geniza, M.; Gibbon, D.; Girard, A.-M.; Haggerty, R.; Higley, K.; Hynes, D.; Lubchenco, J.; McLaughlin, K.; Nieto, F. J.; Noakes, A.; Peterson, M.; Piemonti, A.; Sanders, J.; Tyler, B.; Radniecki, T. Wastewater-Based Epidemiology Predicts COVID-19 Community Prevalence. Res. Sq. 2021. https://doi.org/10.21203/rs.3.rs-690031/v1.

13. McClary-Gutierrez, J. S.; Mattioli, M. C.; Marcenac, P.; Silverman, A. I.; Boehm, A. B.; Bibby, K.; Balliet, M.; de los Reyes, F. L.; Gerrity, D.; Griffith, J. F.; Holden, P. A.; Katehis, D.; Kester, G.; LaCross, N.; Lipp, E. K.; Meiman, J.; Noble, R. T.; Brossard, D.; McLellan, S. L. SARS-CoV-2 Wastewater Surveillance for Public Health Action. Emerg. Infect. Dis. 2021, 27 (9), e210753. https://doi.org/10.3201/eid2709.210753.

14. H. Holm, R.; Mukherjee, A.; P. Rai, J.; A. Yeager, R.; Talley, D.; N. Rai, S.; Bhatnagar, A.; Smith, T. SARS-CoV-2 RNA Abundance in Wastewater as a Function of Distinct Urban Sewershed Size. Environ. Sci. Water Res. Technol. 2022, 8 (4), 807–819. https://doi.org/10.1039/D1EW00672J.

15. Scassa, T.; Robinson, P.; Mosoff, R. The Datafication of Wastewater:: Legal, Ethical and Civic Considerations. Technol. Regul. 2022, 2022, 23–35. https://doi.org/10.26116/techreg.2022.003.

16. World Health Organization. WHO Guidelines on Ethical Issues in Public Health Surveillance; World Health Organization: Geneva, 2017.

17. Wu, F.; Zhang, J.; Xiao, A.; Gu, X.; Lee, W. L.; Armas, F.; Kauffman, K.; Hanage, W.; Matus, M.; Ghaeli, N.; Endo, N.; Duvallet, C.; Poyet, M.; Moniz, K.; Washburne, A. D.; Erickson, T. B.; Chai, P. R.; Thompson, J.; Alm, E. J. SARS-CoV-2 Titers in Wastewater Are Higher than Expected from Clinically Confirmed Cases. mSystems 2020, 5 (4), e00614–20. https://doi.org/10.1128/mSystems.00614-20.

18. Jacobs, D.; McDaniel, T.; Varsani, A.; Halden, R. U.; Forrest, S.; Lee, H. Wastewater Monitoring Raises Privacy and Ethical Considerations. *IEEE Trans*. Technol. Soc. 2021, 2 (3), 116–121. https://doi.org/10.1109/TTS.2021.3073886.

19. Gable, L.; Ram, N.; Ram, J. L. Legal and Ethical Implications of Wastewater Monitoring of SARS-CoV-2 for COVID-19 Surveillance. J. Law Biosci. 2020, 7 (1), lsaa039. https://doi.org/10.1093/jlb/lsaa039.

20. Hall, W.; Prichard, J.; Kirkbride, P.; Bruno, R.; Thai, P. K.; Gartner, C.; Lai, F. Y.; Ort, C.; Mueller, J. F. An Analysis of Ethical Issues in Using Wastewater Analysis to Monitor Illicit Drug Use. Addiction 2012, 107 (10), 1767–1773. https://doi.org/10.1111/j.1360-0443.2012.03887.x.

21. Sims, N.; Kasprzyk-Hordern, B. Future Perspectives of Wastewater-Based Epidemiology: Monitoring Infectious Disease Spread and Resistance to the Community Level. Environ. Int. 2020, 139, 105689. https://doi.org/10.1016/j.envint.2020.105689.

22. Shultz, J. M.; Cooper, J. L.; Baingana, F.; Oquendo, M. A.; Espinel, Z.; Althouse, B. M.; Marcelin, L. H.; Towers, S.; Espinola, M.; McCoy, C. B.; Mazurik, L.; Wainberg, M. L.; Neria, Y.; Rechkemmer, A. The Role of Fear-Related Behaviors in the 2013–2016 West Africa Ebola Virus Disease Outbreak. Curr. Psychiatry Rep. 2016, 18 (11), 104. https://doi.org/10.1007/s11920-016-0741-y.

23. Price, M.; Trowsdale, S. The Ethics of Wastewater Surveillance for Public Health. J. Hydrol. N. Z. 2022, 61 (1), 59–75. https://doi.org/10.3316/informit.548101814967529.

24. LaJoie, A. S.; Holm, R. H.; Anderson, L. B.; Ness, H. D.; Smith, T. Nationwide Public Perceptions Regarding the Acceptance of Using Wastewater for Community Health Monitoring in the United States. PLOS ONE 2022, 17 (10), e0275075. https://doi.org/10.1371/journal.pone.0275075.

25. Polo, D.; Quintela-Baluja, M.; Corbishley, A.; Jones, D. L.; Singer, A. C.; Graham, D. W.; Romalde, J. L. Making Waves: Wastewater-Based Epidemiology for COVID-19 – Approaches and Challenges for Surveillance and Prediction. Water Res. 2020, 186, 116404. https://doi.org/10.1016/j.watres.2020.116404.

26. Coffman, M. M.; Guest, J. S.; Wolfe, M. K.; Naughton, C. C.; Boehm, A. B.; Vela, J. D.; Carrera, J. S. Preventing Scientific and Ethical Misuse of Wastewater Surveillance Data. Environ. Sci. Technol. 2021, 55 (17), 11473–11475. https://doi.org/10.1021/acs.est.1c04325.

27. Ram, N.; Shuster, W.; Gable, L.; Ram, J. L. Ethical and Legal Wastewater Surveillance. Science 2023, 379 (6633), 652–652. https://doi.org/10.1126/science.adg7147.

28. Holm, R. H.; Brick, J. M.; Amraotkar, A. R.; Hart, J. L.; Mukherjee, A.; Zeigler, J.; Bushau-Sprinkle, A. M.; Anderson, L. B.; Walker, K. L.; Talley, D.; Keith, R. J.; Rai, S. N.; Palmer, K. E.; Bhatnagar, A.; Smith, T. Public Awareness of and Support for the Use of Wastewater for SARS-CoV-2 Monitoring: A Community Survey in Louisville, Kentucky. ACS EST Water 2022. https://doi.org/10.1021/acsestwater.1c00405.

29. Thompson, J. R.; Nancharaiah, Y. V.; Gu, X.; Lee, W. L.; Rajal, V. B.; Haines, M. B.; Girones, R.; Ng, L. C.; Alm, E. J.; Wuertz, S. Making Waves: Wastewater Surveillance of SARS-CoV-2 for Population-Based Health Management. Water Res. 2020, 184, 116181. https://doi.org/10.1016/j.watres.2020.116181.

30. Cooper, B.; Donner, E.; Crase, L.; Robertson, H.; Carter, D.; Short, M.; Drigo, B.; Leder, K.; Roiko, A.; Fielding, K. Maintaining a Social License to Operate for Wastewater-Based Monitoring: The Case of Managing Infectious Disease and the COVID-19 Pandemic. J. Environ. Manage. 2022, 320, 115819. https://doi.org/10.1016/j.jenvman.2022.115819.

31. Bowes, D. A.; Driver, E. M.; Halden, R. U. A Framework for Wastewater Sample Collection from a Sewage Cleanout to Inform Building-Scale Wastewater-Based Epidemiology Studies. Sci. Total Environ. 2022, 836, 155576. https://doi.org/10.1016/j.scitotenv.2022.155576.

32. Hrudey, S. E.; Silva, D. S.; Shelley, J.; Pons, W.; Isaac-Renton, J.; Chik, A. H.-S.; Conant, B. Ethics Guidance for Environmental Scientists Engaged in Surveillance of Wastewater for SARS-CoV-2. Environ. Sci. Technol. 2021, 55 (13), 8484–8491. https://doi.org/10.1021/acs.est.1c00308.

33. Manning, S.; Walton, M. Ethical and Responsible Development of Wastewater-Based Epidemiology Technologies; report; Institute of Environmental Science and Research, 2021. https://doi.org/10.26091/ESRNZ.16825243.v1.

34. Prichard, J.; Hall, W.; Zuccato, E.; Voogt, P.; Voulvoulis, N.; Kummerer, K.; Kasprzyk-Hordern, B.; Barbato, A.; Parabiaghi, A.; Hernandez, F.; others. Ethical Research Guidelines for Wastewater-Based Epidemiology and Related Fields. Sew. Anal. Core Group Eur. SCORE 2016.

35. Prichard, J.; Hall, W.; de Voogt, P.; Zuccato, E. Sewage Epidemiology and Illicit Drug Research: The Development of Ethical Research Guidelines. Sci. Total Environ. 2014, 472, 550–555. https://doi.org/10.1016/j.scitotenv.2013.11.039.

36. Savulescu, J. Editorial: Two Deaths and Two Lessons: Is It Time to Review the Structure and Function of Research Ethics Committees? J. Med. Ethics 2002, 28 (1), 1–2.

37. Marx, G. T. Ethics for the New Surveillance. Inf. Soc. 1998, 14 (3), 171–185. https://doi.org/10.1080/019722498128809.

38. Wong, J. C. C.; Tan, J.; Lim, Y. X.; Arivalan, S.; Hapuarachchi, H. C.; Mailepessov, D.; Griffiths, J.; Jayarajah, P.; Setoh, Y. X.; Tien, W. P.; Low, S. L.; Koo, C.; Yenamandra, S. P.; Kong, M.; Lee, V. J. M.; Ng, L. C. Non-Intrusive Wastewater Surveillance for Monitoring of a Residential Building for COVID-19 Cases. Sci. Total Environ. 2021, 786, 147419. https://doi.org/10.1016/j.scitotenv.2021.147419.

39. Kirby, A. E.; Walters, M. S.; Jennings, W. C.; Fugitt, R.; LaCross, N.; Mattioli, M.; Marsh, Z. A.; Roberts, V. A.; Mercante, J. W.; Yoder, J.; Hill, V. R. Using Wastewater Surveillance Data to Support the COVID-19 Response — United States, 2020–2021. Morb. Mortal. Wkly. Rep. 2021, 70 (36), 1242–1244. https://doi.org/10.15585/mmwr.mm7036a2.

40. Gupta, U. C. Informed Consent in Clinical Research: Revisiting Few Concepts and Areas. Perspect. Clin. Res. 2013, 4 (1), 26–32. https://doi.org/10.4103/2229-3485.106373.

41. Lee, L. M. Health Information in the Background: Justifying Public Health Surveillance Without Patient Consent. In Emerging Pervasive Information and Communication Technologies (PICT): Ethical Challenges, Opportunities and Safeguards; Pimple, K. D., Ed.; Law, Governance and Technology Series; Springer Netherlands: Dordrecht, 2014; pp 39–53. https://doi.org/10.1007/978-94-007-6833-8_3.

42. Wright, J.; Driver, E. M.; Bowes, D. A.; Johnston, B.; Halden, R. U. Comparison of High-Frequency in-Pipe SARS-CoV-2 Wastewater-Based Surveillance to Concurrent COVID-19 Random Clinical Testing on a Public U.S. University Campus. Sci. Total Environ. 2022, 820, 152877. https://doi.org/10.1016/j.scitotenv.2021.152877.

43. Betancourt, W. Q.; Schmitz, B. W.; Innes, G. K.; Prasek, S. M.; Pogreba Brown, K. M.; Stark, E. R.; Foster, A. R.; Sprissler, R. S.; Harris, D. T.; Sherchan, S. P.; Gerba, C. P.; Pepper, I. L. COVID-19 Containment on a College Campus via Wastewater-Based Epidemiology, Targeted Clinical Testing and an Intervention. Sci. Total Environ. 2021, 779, 146408. https://doi.org/10.1016/j.scitotenv.2021.146408.

44. Schmitz, B. W.; Innes, G. K.; Prasek, S. M.; Betancourt, W. Q.; Stark, E. R.; Foster, A. R.; Abraham, A. G.; Gerba, C. P.; Pepper, I. L. Enumerating Asymptomatic COVID-19 Cases and Estimating SARS-CoV-2 Fecal Shedding Rates via Wastewater-Based Epidemiology. Sci. Total Environ. 2021, 801, 149794. https://doi.org/10.1016/j.scitotenv.2021.149794.

45. Reeves, K.; Liebig, J.; Feula, A.; Saldi, T.; Lasda, E.; Johnson, W.; Lilienfeld, J.; Maggi, J.; Pulley, K.; Wilkerson, P. J.; Real, B.; Zak, G.; Davis, J.; Fink, M.; Gonzales, P.; Hager, C.; Ozeroff, C.; Tat, K.; Alkire, M.; Butler, C.; Coe, E.; Darby, J.; Freeman, N.; Heuer, H.; Jones, J. R.; Karr, M.; Key, S.; Maxwell, K.; Nelson, L.; Saldana, E.; Shea, R.; Salveson, L.; Tomlinson, K.; Vargas-Barriga, J.; Vigil, B.; Brisson, G.; Parker, R.; Leinwand, L. A.; Bjorkman, K.; Mansfeldt, C. High-Resolution within-Sewer SARS-CoV-2 Surveillance Facilitates Informed Intervention. Water Res. 2021, 204, 117613. https://doi.org/10.1016/j.watres.2021.117613.

46. Brooks, Y. M.; Gryskwicz, B.; Sheehan, S.; Piers, S.; Mahale, P.; McNeil, S.; Chase, J.; Webber, D.; Borys, D.; Hilton, M.; Robinson, D.; Sears, S.; Smith, E.; Lesher, E. K.; Wilson, R.; Goodwin, M.; Pardales, M. Detection of SARS-CoV-2 in Wastewater at Residential College, Maine, USA, August–November 2020. Emerg. Infect. Dis. 2021, 27 (12), 3111–3114. https://doi.org/10.3201/eid2712.211199.

47. Coxon, S. E.; Hopley, T.; Gilpin, B. J. Exploring Opportunities for Sewage Testing on Cargo Ships as a Tool to Screen Seafarers for COVID-19. J. Hydrol. N. Z. 2022, 61 (1), 5–30. https://doi.org/10.3316/informit.548045916053754.

48. Ahmed, W.; Bivins, A.; Simpson, S. L.; Bertsch, P. M.; Ehret, J.; Hosegood, I.; Metcalfe, S. S.; Smith, W. J. M.; Thomas, K. V.; Tynan, J.; Mueller, J. F. Wastewater Surveillance Demonstrates High Predictive Value for COVID-19 Infection on Board Repatriation Flights to Australia. Environ. Int. 2022, 158, 106938. https://doi.org/10.1016/j.envint.2021.106938.

49. Safford, H. R.; Shapiro, K.; Bischel, H. N. Wastewater Analysis Can Be a Powerful Public Health Tool—If It’s Done Sensibly. Proc. Natl. Acad. Sci. 2022, 119 (6), e2119600119. https://doi.org/10.1073/pnas.2119600119.

50. Acosta, N.; Bautista, M. A.; Hollman, J.; McCalder, J.; Beaudet, A. B.; Man, L.; Waddell, B. J.; Chen, J.; Li, C.; Kuzma, D.; Bhatnagar, S.; Leal, J.; Meddings, J.; Hu, J.; Cabaj, J. L.; Ruecker, N. J.; Naugler, C.; Pillai, D. R.; Achari, G.; Ryan, M. C.; Conly, J. M.; Frankowski, K.; Hubert, C. R.; Parkins, M. D. A Multicenter Study Investigating SARS-CoV-2 in Tertiary-Care Hospital Wastewater. Viral Burden Correlates with Increasing Hospitalized Cases as Well as Hospital-Associated Transmissions and Outbreaks. Water Res. 2021, 201, 117369. https://doi.org/10.1016/j.watres.2021.117369.

51. Ahmed, W.; Angel, N.; Edson, J.; Bibby, K.; Bivins, A.; O’Brien, J. W.; Choi, P. M.; Kitajima, M.; Simpson, S. L.; Li, J.; Tscharke, B.; Verhagen, R.; Smith, W. J. M.; Zaugg, J.; Dierens, L.; Hugenholtz, P.; Thomas, K. V.; Mueller, J. F. First Confirmed Detection of SARS-CoV-2 in Untreated Wastewater in Australia: A Proof of Concept for the Wastewater Surveillance of COVID-19 in the Community. Sci. Total Environ. 2020, 728, 138764. https://doi.org/10.1016/j.scitotenv.2020.138764.

52. Ahmed, F.; Islam, Md. A.; Kumar, M.; Hossain, M.; Bhattacharya, P.; Islam, Md. T.; Hossen, F.; Hossain, Md. S.; Islam, Md. S.; Uddin, Md. M.; Islam, Md. N.; Bahadur, N. M.; Didar-ul-Alam, Md.; Reza, H. M.; Jakariya, Md. First Detection of SARS-CoV-2 Genetic Material in the Vicinity of COVID-19 Isolation Centre in Bangladesh: Variation along the Sewer Network. Sci. Total Environ. 2021, 776, 145724. https://doi.org/10.1016/j.scitotenv.2021.145724.

53. Albastaki, A.; Naji, M.; Lootah, R.; Almeheiri, R.; Almulla, H.; Almarri, I.; Alreyami, A.; Aden, A.; Alghafri, R. First Confirmed Detection of SARS-COV-2 in Untreated Municipal and Aircraft Wastewater in Dubai, UAE: The Use of Wastewater Based Epidemiology as an Early Warning Tool to Monitor the Prevalence of COVID-19. Sci. Total Environ. 2021, 760, 143350. https://doi.org/10.1016/j.scitotenv.2020.143350.

54. Arora, S.; Nag, A.; Sethi, J.; Rajvanshi, J.; Saxena, S.; Shrivastava, S. K.; Gupta, A. B. Sewage Surveillance for the Presence of SARS-CoV-2 Genome as a Useful Wastewater Based Epidemiology (WBE) Tracking Tool in India. Water Sci. Technol. J. Int. Assoc. Water Pollut. Res. 2020, 82 (12), 2823–2836. https://doi.org/10.2166/wst.2020.540.

55. Barrios, R. E.; Lim, C.; Kelley, M. S.; Li, X. SARS-CoV-2 Concentrations in a Wastewater Collection System Indicated Potential COVID-19 Hotspots at the Zip Code Level. Sci. Total Environ. 2021, 800, 149480. https://doi.org/10.1016/j.scitotenv.2021.149480.

56. Bivins, A.; Bibby, K. Wastewater Surveillance during Mass COVID-19 Vaccination on a College Campus. Environ. Sci. Technol. Lett. 2021, 8 (9), 792–798. https://doi.org/10.1021/acs.estlett.1c00519.

57. Bivins, A.; Lott, M.; Shaffer, M.; Wu, Z.; North, D.; K. Lipp, E.; Bibby, K. Building-Level Wastewater Surveillance Using Tampon Swabs and RT-LAMP for Rapid SARS-CoV-2 RNA Detection. Environ. Sci. Water Res. Technol. 2022, 8 (1), 173–183. https://doi.org/10.1039/D1EW00496D.

58. Black, J.; Aung, P.; Nolan, M.; Roney, E.; Poon, R.; Hennessy, D.; Crosbie, N. D.; Deere, D.; Jex, A. R.; John, N.; Baker, L.; Scales, P. J.; Usher, S. P.; McCarthy, D. T.; Schang, C.; Schmidt, J.; Myers, S.; Begue, N.; Kaucner, C.; Thorley, B.; Druce, J.; Monis, P.; Lau, M.; Sarkis, S. Epidemiological Evaluation of Sewage Surveillance as a Tool to Detect the Presence of COVID-19 Cases in a Low Case Load Setting. Sci. Total Environ. 2021, 786, 147469. https://doi.org/10.1016/j.scitotenv.2021.147469.

59. Carrillo-Reyes, J.; Barragán-Trinidad, M.; Buitrón, G. Surveillance of SARS-CoV-2 in Sewage and Wastewater Treatment Plants in Mexico. J. Water Process Eng. 2021, 40, 101815. https://doi.org/10.1016/j.jwpe.2020.101815.

60. Chakraborty, P.; Pasupuleti, M.; Jai Shankar, M. R.; Bharat, G. K.; Krishnasamy, S.; Dasgupta, S. C.; Sarkar, S. K.; Jones, K. C. First Surveillance of SARS-CoV-2 and Organic Tracers in Community Wastewater during Post Lockdown in Chennai, South India: Methods, Occurrence and Concurrence. Sci. Total Environ. 2021, 778, 146252. https://doi.org/10.1016/j.scitotenv.2021.146252.

61. Colosi, L. M.; Barry, K. E.; Kotay, S. M.; Porter, M. D.; Poulter, M. D.; Ratliff, C.; Simmons, W.; Steinberg, L. I.; Wilson, D. D.; Morse, R.; Zmick, P.; Mathers, A. J. Development of Wastewater Pooled Surveillance of Severe Acute Respiratory Syndrome Coronavirus 2 (SARS-CoV-2) from Congregate Living Settings. Appl. Environ. Microbiol. 2021. https://doi.org/10.1128/AEM.00433-21.

62. Corchis-Scott, R.; Geng, Q.; Seth, R.; Ray, R.; Beg, M.; Biswas, N.; Charron, L.; Drouillard, K. D.; D’Souza, R.; Heath, D. D.; Houser, C.; Lawal, F.; McGinlay, J.; Menard, S. L.; Porter, L. A.; Rawlings, D.; Scholl, M. L.; Siu, K. W. M.; Tong, Y.; Weisener, C. G.; Wilhelm, S. W.; McKay, R. M. L. Averting an Outbreak of SARS-CoV-2 in a University Residence Hall through Wastewater Surveillance. Microbiol. Spectr. 2021. https://doi.org/10.1128/Spectrum.00792-21.

63. Crowe, J.; Schnaubelt, A. T.; SchmidtBonne, S.; Angell, K.; Bai, J.; Eske, T.; Nicklin, M.; Pratt, C.; White, B.; Crotts-Hannibal, B.; Staffend, N.; Herrera, V.; Cobb, J.; Conner, J.; Carstens, J.; Tempero, J.; Bouda, L.; Ray, M.; Lawler, J. V.; Campbell, W. S.; Lowe, J.- M.; Santarpia, J.; Bartelt-Hunt, S.; Wiley, M.; Brett-Major, D.; Logan, C.; Broadhurst, M. J. Assessment of a Program for SARS-CoV-2 Screening and Environmental Monitoring in an Urban Public School District. *JAMA Netw*. Open 2021, 4 (9), e2126447. https://doi.org/10.1001/jamanetworkopen.2021.26447.

64. Daigle, J.; Racher, K.; Hazenberg, J.; Yeoman, A.; Hannah, H.; Duong, D.; Mohammed, U.; Spreitzer, D.; Gregorchuk, B. S. J.; Head, B. M.; Meyers, A. F. A.; Sandstrom, P. A.; Nichani, A.; Brooks, J. I.; Mulvey, M. R.; Mangat, C. S.; Becker, M. G. A Sensitive and Rapid Wastewater Test for SARS-COV-2 and Its Use for the Early Detection of a Cluster of Cases in a Remote Community. medRxiv 2021, 2021.08.13.21262039. https://doi.org/10.1101/2021.08.13.21262039.

65. Fahrenfeld, N. L.; Morales Medina, W. R.; D’Elia, S.; Modica, M.; Ruiz, A.; McLane, M. Comparison of Residential Dormitory COVID-19 Monitoring via Weekly Saliva Testing and Sewage Monitoring. Sci. Total Environ. 2021, 151947. https://doi.org/10.1016/j.scitotenv.2021.151947.

66. Fongaro, G.; Rogovski, P.; Savi, B. P.; Cadamuro, R. D.; Pereira, J. V. F.; Anna, I. H. S.; Rodrigues, I. H.; Souza, D. S. M.; Saravia, E. G. T.; Rodríguez-Lázaro, D.; da Silva Lanna, M. C. SARS-CoV-2 in Human Sewage and River Water from a Remote and Vulnerable Area as a Surveillance Tool in Brazil. Food Environ. Virol. 2021. https://doi.org/10.1007/s12560-021-09487-9.

67. Galani, A.; Aalizadeh, R.; Kostakis, M.; Markou, A.; Alygizakis, N.; Lytras, T.; Adamopoulos, P. G.; Peccia, J.; Thompson, D. C.; Kontou, A.; Karagiannidis, A.; Lianidou, E. S.; Avgeris, M.; Paraskevis, D.; Tsiodras, S.; Scorilas, A.; Vasiliou, V.; Dimopoulos, M.-A.; Thomaidis, N. S. SARS-CoV-2 Wastewater Surveillance Data Can Predict Hospitalizations and ICU Admissions. Sci. Total Environ. 2022, 804, 150151. https://doi.org/10.1016/j.scitotenv.2021.150151.

68. Gerrity, D.; Papp, K.; Stoker, M.; Sims, A.; Frehner, W. Early-Pandemic Wastewater Surveillance of SARS-CoV-2 in Southern Nevada: Methodology, Occurrence, and Incidence/Prevalence Considerations. Water Res. X 2021, 10, 100086. https://doi.org/10.1016/j.wroa.2020.100086.

69. Gibas, C.; Lambirth, K.; Mittal, N.; Juel, M. A. I.; Barua, V. B.; Roppolo Brazell, L.; Hinton, K.; Lontai, J.; Stark, N.; Young, I.; Quach, C.; Russ, M.; Kauer, J.; Nicolosi, B.; Chen, D.; Akella, S.; Tang, W.; Schlueter, J.; Munir, M. Implementing Building-Level SARS-CoV-2 Wastewater Surveillance on a University Campus. Sci. Total Environ. 2021, 782, 146749. https://doi.org/10.1016/j.scitotenv.2021.146749.

70. Giraud-Billoud, M.; Cuervo, P.; Altamirano, J. C.; Pizarro, M.; Aranibar, J. N.; Catapano, A.; Cuello, H.; Masachessi, G.; Vega, I. A. Monitoring of SARS-CoV-2 RNA in Wastewater as an Epidemiological Surveillance Tool in Mendoza, Argentina. Sci. Total Environ. 2021, 796, 148887. https://doi.org/10.1016/j.scitotenv.2021.148887.

71. Godinez, A.; Hill, D.; Dandaraw, B.; Green, H.; Kilaru, P.; Middleton, F.; Run, S.; Kmush, B. L.; Larsen, D. A. High Sensitivity and Specificity of Dormitory-Level Wastewater Surveillance for COVID-19 during Fall Semester 2020 at Syracuse University, New York. Int. J. Environ. Res. Public. Health 2022, 19 (8), 4851. https://doi.org/10.3390/ijerph19084851.

72. Gonçalves, J.; Koritnik, T.; Mioč, V.; Trkov, M.; Bolješič, M.; Berginc, N.; Prosenc, K.; Kotar, T.; Paragi, M. Detection of SARS-CoV-2 RNA in Hospital Wastewater from a Low COVID-19 Disease Prevalence Area. Sci. Total Environ. 2021, 755, 143226. https://doi.org/10.1016/j.scitotenv.2020.143226.

73. Haak, L.; Delic, B.; Li, L.; Guarin, T.; Mazurowski, L.; Dastjerdi, N. G.; Dewan, A.; Pagilla, K. Spatial and Temporal Variability and Data Bias in Wastewater Surveillance of SARS-CoV-2 in a Sewer System. Sci. Total Environ. 2022, 805, 150390. https://doi.org/10.1016/j.scitotenv.2021.150390.

74. Hasan, S. W.; Ibrahim, Y.; Daou, M.; Kannout, H.; Jan, N.; Lopes, A.; Alsafar, H.; Yousef, A. F. Detection and Quantification of SARS-CoV-2 RNA in Wastewater and Treated Effluents: Surveillance of COVID-19 Epidemic in the United Arab Emirates. Sci. Total Environ. 2021, 764, 142929–142929. https://doi.org/10.1016/j.scitotenv.2020.142929.

75. Hong, P.-Y.; Rachmadi, A. T.; Mantilla-Calderon, D.; Alkahtani, M.; Bashawri, Y. M.; Al Qarni, H.; O’Reilly, K. M.; Zhou, J. Estimating the Minimum Number of SARS-CoV-2 Infected Cases Needed to Detect Viral RNA in Wastewater: To What Extent of the Outbreak Can Surveillance of Wastewater Tell Us? Environ. Res. 2021, 195, 110748. https://doi.org/10.1016/j.envres.2021.110748.

76. Juel, M. A. I.; Stark, N.; Nicolosi, B.; Lontai, J.; Lambirth, K.; Schlueter, J.; Gibas, C.; Munir, M. Performance Evaluation of Virus Concentration Methods for Implementing SARS-CoV-2 Wastewater Based Epidemiology Emphasizing Quick Data Turnaround. Sci. Total Environ. 2021, 801, 149656. https://doi.org/10.1016/j.scitotenv.2021.149656.

77. Karami, C.; Dargahi, A.; Vosoughi, M.; Normohammadi, A.; Jeddi, F.; Asghariazar, V.; Mokhtari, A.; Sedigh, A.; Zandian, H.; Alighadri, M. SARS-CoV-2 in Municipal Wastewater Treatment Plant, Collection Network, and Hospital Wastewater. Environ. Sci. Pollut. Res. 2021. https://doi.org/10.1007/s11356-021-15374-4.

78. Karthikeyan, S.; Nguyen, A.; McDonald, D.; Zong, Y.; Ronquillo, N.; Ren, J.; Zou, J.; Farmer, S.; Humphrey, G.; Henderson, D.; Javidi, T.; Messer, K.; Anderson, C.; Schooley, R.; Martin, N. K.; Knight, R. Rapid, Large-Scale Wastewater Surveillance and Automated Reporting System Enable Early Detection of Nearly 85% of COVID-19 Cases on a University Campus. mSystems 2021. https://doi.org/10.1128/mSystems.00793-21.

79. Kitamura, K.; Sadamasu, K.; Muramatsu, M.; Yoshida, H. Efficient Detection of SARS-CoV-2 RNA in the Solid Fraction of Wastewater. Sci. Total Environ. 2021, 763, 144587. https://doi.org/10.1016/j.scitotenv.2020.144587.

80. Koureas, M.; Amoutzias, G. D.; Vontas, A.; Kyritsi, M.; Pinaka, O.; Papakonstantinou, A.; Dadouli, K.; Hatzinikou, M.; Koutsolioutsou, A.; Mouchtouri, V. A.; Speletas, M.; Tsiodras, S.; Hadjichristodoulou, C. Wastewater Monitoring as a Supplementary Surveillance Tool for Capturing SARS-COV-2 Community Spread. A Case Study in Two Greek Municipalities. Environ. Res. 2021, 200, 111749. https://doi.org/10.1016/j.envres.2021.111749.

81. Kumar, M.; Patel, A. K.; Shah, A. V.; Raval, J.; Rajpara, N.; Joshi, M.; Joshi, C. G. First Proof of the Capability of Wastewater Surveillance for COVID-19 in India through Detection of Genetic Material of SARS-CoV-2. Sci. Total Environ. 2020, 746, 141326. https://doi.org/10.1016/j.scitotenv.2020.141326.

82. Landstrom, M.; Braun, E.; Larson, E.; Miller, M.; Holm, G. H. Efficacy of SARS-CoV-2 Wastewater Surveillance for Detection of COVID-19 at a Residential Private College. medRxiv September 22, 2021, p 2021.09.15.21263338. https://doi.org/10.1101/2021.09.15.21263338.

83. Li, B.; Di, D. Y. W.; Saingam, P.; Jeon, M. K.; Yan, T. Fine-Scale Temporal Dynamics of SARS-CoV-2 RNA Abundance in Wastewater during A COVID-19 Lockdown. Water Res. 2021, 197, 117093. https://doi.org/10.1016/j.watres.2021.117093.

84. Melvin, R. G.; Hendrickson, E. N.; Chaudhry, N.; Georgewill, O.; Freese, R.; Schacker, T. W.; Simmons, G. E. A Novel Wastewater-Based Epidemiology Indexing Method Predicts SARS-CoV-2 Disease Prevalence across Treatment Facilities in Metropolitan and Regional Populations. Sci. Rep. 2021, 11 (1), 21368. https://doi.org/10.1038/s41598-021-00853-y.

85. Mota, C. R.; Bressani-Ribeiro, T.; Araújo, J. C.; Leal, C. D.; Leroy-Freitas, D.; Machado, E. C.; Espinosa, M. F.; Fernandes, L.; Leão, T. L.; Chamhum-Silva, L.; Azevedo, L.; Morandi, T.; Freitas, G. T. O.; Costa, M. S.; Carvalho, B. O.; Reis, M. T. P.; Melo, M. C.; Ayrimoraes, S. R.; Chernicharo, C. A. L. Assessing Spatial Distribution of COVID-19 Prevalence in Brazil Using Decentralised Sewage Monitoring. Water Res. 2021, 202, 117388–117388. https://doi.org/10.1016/j.watres.2021.117388.

86. Nemudryi, A.; Nemudraia, A.; Wiegand, T.; Surya, K.; Buyukyoruk, M.; Cicha, C.; Vanderwood, K. K.; Wilkinson, R.; Wiedenheft, B. Temporal Detection and Phylogenetic Assessment of SARS-CoV-2 in Municipal Wastewater. Cell Rep. Med. 2020, 1 (6), 100098. https://doi.org/10.1016/j.xcrm.2020.100098.

87. Prado, T.; Fumian, T. M.; Mannarino, C. F.; Resende, P. C.; Motta, F. C.; Eppinghaus, A. L. F.; Chagas do Vale, V. H.; Braz, R. M. S.; de Andrade, J. da S. R.; Maranhão, A. G.; Miagostovich, M. P. Wastewater-Based Epidemiology as a Useful Tool to Track SARS-CoV-2 and Support Public Health Policies at Municipal Level in Brazil. Water Res. 2021, 191, 116810–116810. https://doi.org/10.1016/j.watres.2021.116810.

88. Rodríguez Rasero, F. J.; Moya Ruano, L. A.; Rasero Del Real, P.; Cuberos Gómez, L.; Lorusso, N. Associations between SARS-CoV-2 RNA Concentrations in Wastewater and COVID-19 Rates in Days after Sampling in Small Urban Areas of Seville: A Time Series Study. Sci. Total Environ. 2022, 806, 150573. https://doi.org/10.1016/j.scitotenv.2021.150573.

89. Rios, G.; Lacoux, C.; Leclercq, V.; Diamant, A.; Lebrigand, K.; Lazuka, A.; Soyeux, E.; Lacroix, S.; Fassy, J.; Couesnon, A.; Thiery, R.; Mari, B.; Pradier, C.; Waldmann, R.; Barbry, P. Monitoring SARS-CoV-2 Variants Alterations in Nice Neighborhoods by Wastewater Nanopore Sequencing. Lancet Reg. Health - Eur. 2021, 10, 100202. https://doi.org/10.1016/j.lanepe.2021.100202.

90. Scott, L. C.; Aubee, A.; Babahaji, L.; Vigil, K.; Tims, S.; Aw, T. G. Targeted Wastewater Surveillance of SARS-CoV-2 on a University Campus for COVID-19 Outbreak Detection and Mitigation. Environ. Res. 2021, 200, 111374. https://doi.org/10.1016/j.envres.2021.111374.

91. Sherchan, S. P.; Shahin, S.; Ward, L. M.; Tandukar, S.; Aw, T. G.; Schmitz, B.; Ahmed, W.; Kitajima, M. First Detection of SARS-CoV-2 RNA in Wastewater in North America: A Study in Louisiana, USA. Sci. Total Environ. 2020, 743, 140621. https://doi.org/10.1016/j.scitotenv.2020.140621.

92. Spurbeck, R. R.; Minard-Smith, A.; Catlin, L. Feasibility of Neighborhood and Building Scale Wastewater-Based Genomic Epidemiology for Pathogen Surveillance. Sci. Total Environ. 2021, 789, 147829–147829. https://doi.org/10.1016/j.scitotenv.2021.147829.

93. Sweetapple, C.; Melville-Shreeve, P.; Chen, A. S.; Grimsley, J. M. S.; Bunce, J. T.; Gaze, W.; Fielding, S.; Wade, M. J. Building Knowledge of University Campus Population Dynamics to Enhance Near-to-Source Sewage Surveillance for SARS-CoV-2 Detection. Sci. Total Environ. 2022, 806, 150406. https://doi.org/10.1016/j.scitotenv.2021.150406.

94. Vo, V.; Tillett, R. L.; Chang, C.-L.; Gerrity, D.; Betancourt, W. Q.; Oh, E. C. SARS-CoV-2 Variant Detection at a University Dormitory Using Wastewater Genomic Tools. Sci. Total Environ. 2022, 805, 149930. https://doi.org/10.1016/j.scitotenv.2021.149930.

95. Wang, Y.; Liu, P.; Zhang, H.; Ibaraki, M.; VanTassell, J.; Geith, K.; Cavallo, M.; Kann, R.; Saber, L.; Kraft, C. S.; Lane, M.; Shartar, S.; Moe, C. Early Warning of a COVID-19 Surge on a University Campus Based on Wastewater Surveillance for SARS-CoV-2 at Residence Halls. Sci. Total Environ. 2022, 821, 153291. https://doi.org/10.1016/j.scitotenv.2022.153291.

96. Wurtz, N.; Revol, O.; Jardot, P.; Giraud-Gatineau, A.; Houhamdi, L.; Soumagnac, C.; Annessi, A.; Lacoste, A.; Colson, P.; Aherfi, S.; La Scola, B. Monitoring the Circulation of SARS-CoV-2 Variants by Genomic Analysis of Wastewater in Marseille, South-East France. Pathogens 2021, 10 (8), 1042. https://doi.org/10.3390/pathogens10081042.

97. Xu, X.; Zheng, X.; Li, S.; Lam, N. S.; Wang, Y.; Chu, D. K. W.; Poon, L. L. M.; Tun, H. M.; Peiris, M.; Deng, Y.; Leung, G. M.; Zhang, T. The First Case Study of Wastewater-Based Epidemiology of COVID-19 in Hong Kong. Sci. Total Environ. 2021, 790, 148000. https://doi.org/10.1016/j.scitotenv.2021.148000.

98. Yeager, R.; Holm, R. H.; Saurabh, K.; Fuqua, J. L.; Talley, D.; Bhatnagar, A.; Smith, T. Wastewater Sample Site Selection to Estimate Geographically Resolved Community Prevalence of COVID-19: A Sampling Protocol Perspective. GeoHealth 2021, 5 (7), e2021gh000420. https://doi.org/10.1029/2021gh000420.

99. Zdenkova, K.; Bartackova, J.; Cermakova, E.; Demnerova, K.; Dostalkova, A.; Janda, V.; Novakova, Z.; Rumlova, M.; Ambrozova, J. R.; Skodakova, K.; Swierczkova, I.; Sykora, P.; Vejmelkova, D.; Wanner, J.; Bartacek, J. Monitoring COVID-19 Spread in Prague Local Neighborhoods Based on the Presence of SARS-CoV-2 RNA in Wastewater Collected throughout the Sewer Network. medRxiv 2021, 2021.07.28.21261272. https://doi.org/10.1101/2021.07.28.21261272.

100. Korfmacher, K. S.; Harris, -Lovett Sasha. Invited Perspective: Implementation of Wastewater-Based Surveillance Requires Collaboration, Integration, and Community Engagement. Environ. Health Perspect. 130 (5), 051304. https://doi.org/10.1289/EHP11191.

101. Berkman, B. E.; Mastroianni, A. C.; Jamal, L.; Solis, C.; Taylor, H. A.; Hull, S. C. The Ethics of Repurposing Previously Collected Research Biospecimens in an Infectious Disease Pandemic. Ethics Hum. Res. 2021, 43 (2), 2–18. https://doi.org/10.1002/eahr.500083.

102. Vears, D. F.; Borry, P.; Savulescu, J.; Koplin, J. J. Old Challenges or New Issues? Genetic Health Professionals’ Experiences Obtaining Informed Consent in Diagnostic Genomic Sequencing. AJOB Empir. Bioeth. 2021, 12 (1), 12–23. https://doi.org/10.1080/23294515.2020.1823906.

103. Dupras, C.; Bunnik, E. M. Toward a Framework for Assessing Privacy Risks in Multi-Omic Research and Databases. Am. J. Bioeth. 2021, 21 (12), 46–64. https://doi.org/10.1080/15265161.2020.1863516.

104. Yang, Z.; Xu, G.; Reboud, J.; Kasprzyk-Hordern, B.; Cooper, J. M. Monitoring Genetic Population Biomarkers for Wastewater-Based Epidemiology. Anal. Chem. 2017, 89 (18), 9941–9945. https://doi.org/10.1021/acs.analchem.7b02257.

105. Machado, H.; Silva, S.; Cunha, M. Multiple Views of DNA Surveillance: The Surveilled, the Surveillants and the Academics. In Crime, Security and Surveillance. Effects for the Surveillant and the Surveilled; Boom Eleven Publishers, 2012; pp 179–194.

106. *Wastewater-Based Disease Surveillance for Public Health Action*; National Academies of Sciences, Engineer (WASHINGTON, District of Columbia), Ed.; The National Academies Press: WASHINGTON, 2023.

