## Supplemental Figure 1 for "Structured Ethical Review for Wastewater-Based Testing"

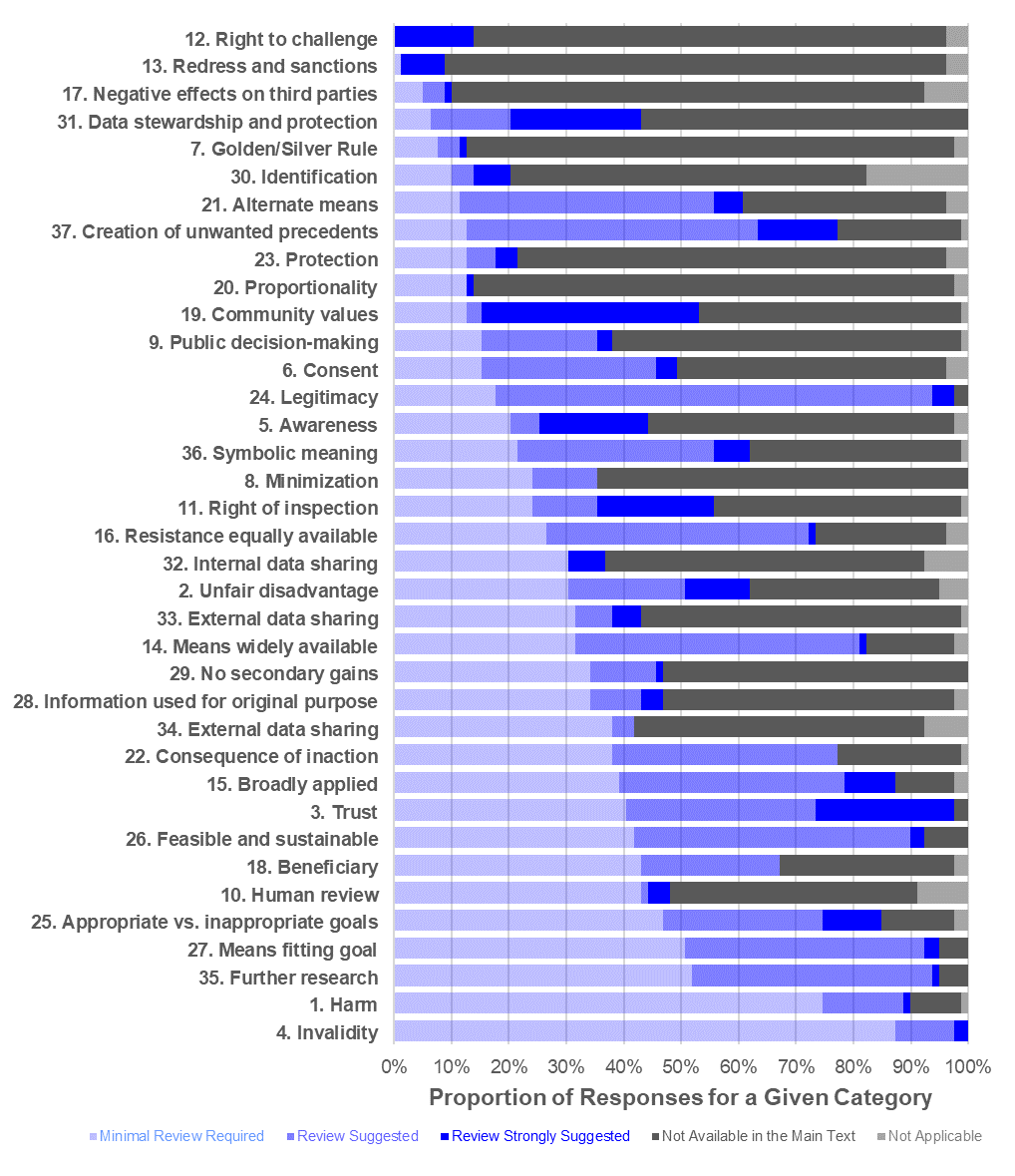

**Supplemental Figure 1.** The distribution of assigned flags (“minimal review required”, “review suggested”, “review strongly suggested”, “not available in the main text”, “not applicable”) for each structured ethical review category represented as a fraction percent of all publications analyzed (n=53) with multiple reviewers providing reports for select individual studies, resulting in more reviews than studies (n=79) ^1,33–37,40–89^. The categories are sorted by ascending proportion of “mnimal review required”.
